# Effect of Vitamin D deficiency on COVID-19 status: A systematic review

**DOI:** 10.1101/2020.12.01.20242313

**Authors:** Pranta Das, Nandeeta Samad, Bright Opoku Ahinkorah, Prince Peprah, Aliu Mohammed, Abdul-Aziz Seidu

## Abstract

**Background:** One major micronutrient known to have a possible protective effect against COVID-19 disease is vitamin D. This systematic review sought to identify and synthesise available evidence to aid the understanding of the possible effect of vitamin D deficiency on COVID-19 status and health outcomes in COVID-19 patients.

**Methods:** Three databases PubMed, ScienceDirect, and Google Scholar were searched systematically to obtain English language journal article published within 1/12/2019 and 3/11/2020. The search consisted of the terms (“Vitamin D,” OR “25□Hydroxyvitamin D,” OR “Low Vitamin D.”) AND (“COVID-19” OR “2019-nCoV” OR “Coronavirus” OR “SARS-CoV-2”) AND (“disease severity” OR “IMV” OR “ICU admission” OR “mortality” OR “hospitalization” OR “infection”). We followed the recommended PRISMA guidelines in executing this study. After going through the screening of the articles, eleven articles were included in the review.

**Findings:** Almost all the included studies reported a positive association between Vitamin D sufficiency and COVID-19 status and health outcomes. Vitamin D deficient patients (< 25 ng/mL) are 5.84 times [aOR=6.84, p=0.01] more likely to die from COVID-19 compared to the vitamin D sufficient people. Another study also found that Vitamin D deficiency is associated with higher risk of death with Hazard ratio (HR) 14.73, p<0.001. Vitamin D deficient (<12 ng/mL) people were 2.2 times [aOR=3.2, p=0.07] more likely to develop severe COVID-19 after adjusting for age, gender, obesity, cardiac disease, and kidney disease compared to the vitamin D sufficient people. One study found that after controlling for confounders, patients with low 25(OH)D (<30 ng/mL) level are more likely [aOR=1.45, p=<0.001] to be COVID-19 infected compared to the patients with 25(OH)D level >=30 ng/mL.

**Conclusion:** Findings from the study included suggest Vitamin D may serve as a mitigating effect for covid-19 infection, severity and mortality. We recommend the need to encourage people to eat foods rich in vitamin D such as fish, red meat, liver and egg yolks whiles at the same time providing vitamin D supplements for individuals with COVID-19 in order to boost their immune systems.

## Introduction

As of 10^th^ November, 2020, approximately 50 million confirmed cases and 1.2 million COVID-19 related deaths had been reported globally (WHO, 2020). There is a seemingly sharp rise in the number of confirmed COVID-19 cases in many countries, in what has been described as the “second wave” of the global pandemic (Looi, 2020). For instance, between September and October, 2020, a number of European countries including Belgium, Germany, France, Spain, Czech Republic and Ireland had reported exponential increases in the daily number of confirmed COVID-19 cases (Looi, 2020). This resurgence of the disease could be attributed to the relaxation of preventive measures such as lockdowns, physical distancing, wearing of nose masks and the general disregard to precautionary behaviours among the populace (Aleta et al., 2020; Leung et al., 2020). Therefore, in the absence of effective pharmacologic therapy and vaccines, it is important to investigate the role of immune boosters such as micronutrients (vitamins and minerals) in mitigating or preventing the adverse effect of the COVID-19 disease (Laird, &Kenny, 2020; Richardson, &Lovegrove, 2020).

One major micronutrient known to have a possible protective or mitigating effect against COVID-19 disease is vitamin D (Ebadi, &Montano-Loza, 2020; Grant et al., 2020; McCartney, &Byrne, 2020), a naturally occurring vitamin in humans, which is mostly produced in the skin after an exposure to ultraviolent radiation from the sun (Di Rosa et al., 2011). Because COVID-19 is associated with immune hyperactivation (Zhong et al., 2020), the protective effect of vitamin D had been attributed to its ability to suppress immune responses to the SARS-CoV-2 virus (Panarese, &Shahini, 2020) and thereby reducing the risk or severity of acute respiratory distress syndrome, a cardinal sign of severe COVID-19 (Grasselli et al., 2020; Zhong et al., 2020). Relatedly, vitamin D deficiency had been associated with increasing risk for immune-mediated and inflammatory disorders including diseases of the respiratory and digestive systems (Di Rosa et al., 2011). For example, many observational studies have shown a significant association between low serum levels of vitamin D and increased risk for acute respiratory tract infections (Ginde et al., 2009; Jolliffe et al., 2013). Additionally, a randomised controlled trial had shown that vitamin D supplementation for patients at high risk of respiratory tract infection reduced symptoms and need for antibiotic therapy (Bergman et a., 2012).

Aside its importance in immune modulation (Bergman et al., 2012; Di Rosa et al., 2011) during early and late phases of SARS-CoV-2 viral infection (Martineau, &Forouhi, 2020), the effect of vitamin D on health outcomes among COVID-19 patients largely remain inconclusive (Ali, 2020; Martineau, &Forouhi, 2020). One major postulate on the possible association of vitamin D deficiency and COVID-19 disease is the high morbidity and mortality recorded among aged population, who are more likely to have lower serum levels of vitamin D (Ebadi, &Montano-Loza, 2020; Laird, &Kenny, 2020). For instance, Ilie et al. (2020) reported that aged population in countries with higher levels of COVID-19 mortality such as Italy and Spain had significantly lower mean serum levels of vitamin D. Additionally, both severe COVID-19 and vitamin D deficiency had been associated with common risk factors such as old age, obesity, and being of Asian or black ethnic descent (Martineau, &Forouhi, 2020). Thus, considering the numerous findings from observational studies on a possible association between vitamin D deficiency and the incidence or severity of COVID-19 disease, we conducted a systematic review with the aim of identifying and synthesising available evidence to aid the understanding of the possible effect of vitamin D deficiency on COVID-19 status and health outcomes in COVID-19 patients.

## Methods

### Search strategy

Three databases PubMed, ScienceDirect, and Google Scholar were searched systematically to obtain English language journal article published within 1/12/2019 and 3/11/2020. The search consisted of the terms (“Vitamin D,” OR “25□Hydroxyvitamin D,” OR “Low Vitamin D.”) AND (“COVID-19” OR “2019-nCoV” OR “Coronavirus” OR “SARS-CoV-2”) AND (“disease severity” OR “IMV” OR “ICU admission” OR “mortality” OR “hospitalization” OR “infection”). More details regarding search strategy are presented in Table S1-S3 in the supplementary file.

### Eligibility Criteria

The studies which dealt with Vitamin D deficiency and assessed outcome of COVID-19 infection, severity, and death among the real-time reverse transcriptase-polymerase chain reaction (RT-PCR) or according to the country specific criteria or laboratory confirmed COVID-19 patients were included. Only peer reviewed journal articles written in English language were included. So unpublished studies, preprints, and articles written in languages except English were excluded. Only studies which are cross-sectional or cohort or case-control study in nature were included. Hence, randomized controlled trails, short communication, letter to the editor, review articles etc. were excluded.

### Data extraction and study quality assessment

After finding pertinent articles by using the inclusion and exclusion criteria, quality of the articles was assessed. Newcastle-Ottawa technique of assessing study quality of cross-sectional study, cohort study and case-control study were used to measure the quality of the cross-sectional, cohort, and case-control studies respectively. Studies with quality score of at least 5, for each study design, were selected for data extraction. Name of the first author, study design, country name, sample size, mean/median age/age interval of the included participants, how vitamin D deficiency was defined, outcome assessed, and main findings were extracted from the included studies.

## Results

### Search results

Through searching the databases; PubMed, Goggle Scholar, and ScienceDirect, 135, 2630, and 9 articles were identified respectively (figure 1). So the titles and abstracts of 2774 articles were initially screened. Through the screening 17 articles were selected for full text screening. After the full text screening of 17 articles 3 articles were excluded due to mismatch of the study design. Then from rest of the eligible articles 3 articles were excluded due to lack of information and duplication. Finally, for qualitative synthesis, 11 articles were included.

**Figure 1:**
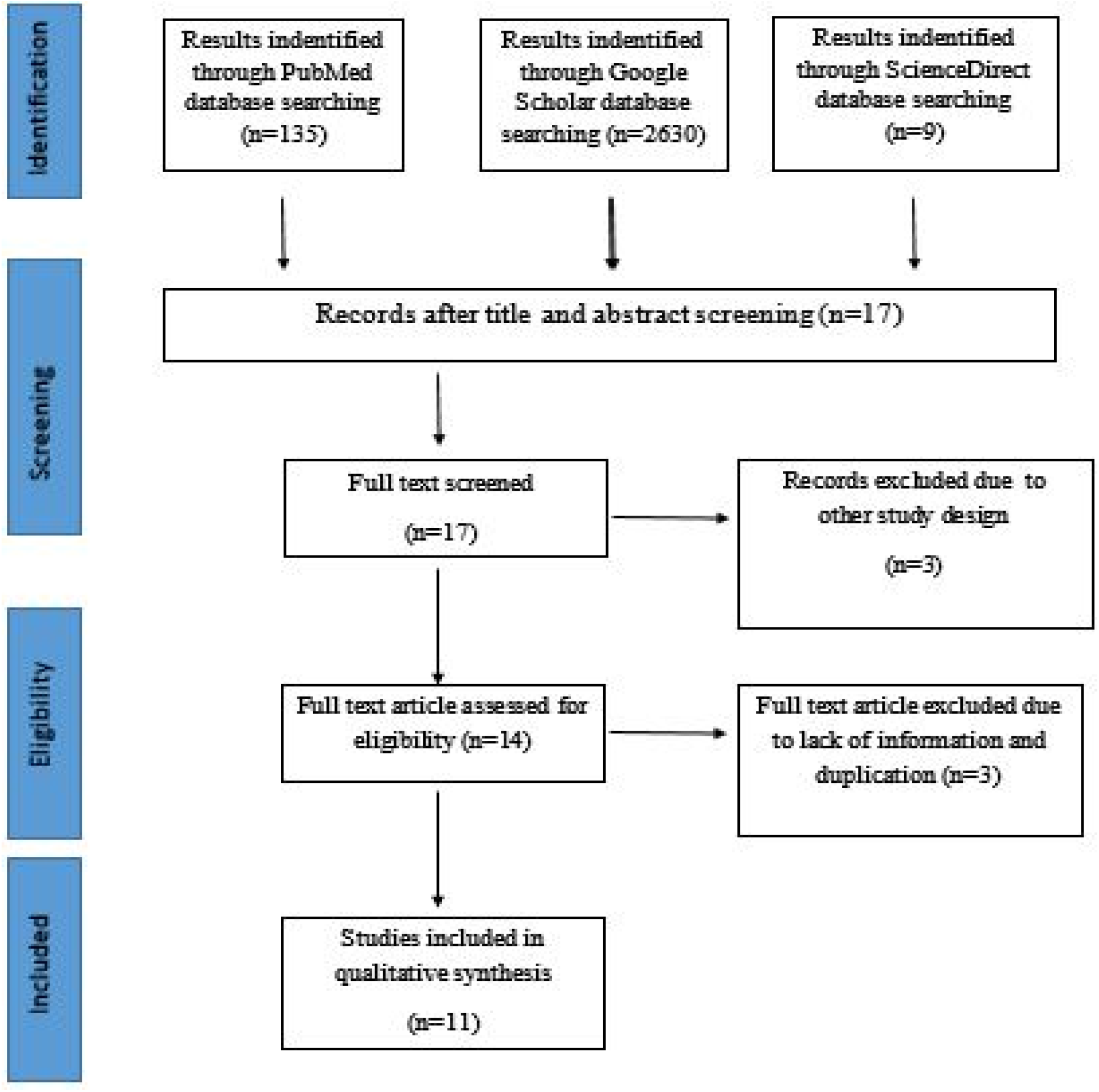
PRISMA flowchart for search strategy and the process of selecting articles

### Study characteristics and main findings

Six studies were cohort studies, four studies were cross-sectional studies and one study was a case-control study among the included studies. Two studies were from USA, UK, Iran each and the rest of the studies were from different countries. Six studies investigated the outcome of COVID-19 infection, three studies investigated severity, and two studies investigated death. Different studies defined vitamin D deficiency differently. All the included studies have moderate to high quality (Table S4-S6 in the supplementary file). More comprehensive characteristics of the studies are presented in Table 1.

**Table 1:** Study characteristics

| Author | Study design | Country | Sample size | Mean/median age /age interval (years) | Vitamin D categories | Outcome assessed |
| --- | --- | --- | --- | --- | --- | --- |
| Abrishami *et al.*, 2020 | Retrospective cohort | Iran | 73 | 55.18 | Deficiency: <25 ng/mL | Death |
| Maghbooli *et al.*, 2020 | Cross-sectional | Iran | 235 | 58.72 | Deficiency: <30 ng/mL | Severity |
| Radujkovic *et al.*, 2020 | Prospective cohort | Germany | 185 | 60 | Deficiency: <12 ng/mL | Death |
| Hastie *et al.*, 2020 | Cross-sectional | UK | 348598 | 38 – 58 | Deficiency: <25 nmol/L | Infection |
| Otros *et al.*, 2020 | Retrospective cohort | Spain | 80 | 50 – 84 | Deficiency: <20 ng/mL | Severity |
| Merzon *et al.*, 2020 | Cross-sectional | Israel | 7807 | 46.17 | Deficiency: <30 ng/ml | Infection |
| Meltzer *et al.*, 2020 | Retrospective cohort | USA | 489 | 49.2 | Deficiency: <20 ng/mL | Infection |
| Ye *et al.*, 2020 | Case-control study | China | 142 | 43 | Deficiency: <50 nmol/L | Severity |
| Kaufman *et al.*, 2020 | Cross-sectional | USA | 191779 | 54 | Deficiency: <20 ng/mL | Infection |
| D'avolio *et al.*, 2020 | Retrospective cohort | Switzerland | 107 | 73 | Not categorized | Infection |
| Baktash *et al.*, 2020 | Prospective cohort study | UK | 105 | 81.29 | Deficiency: <=30 nmol/L | Infection |

### Findings from all the studies

The study by Abrishami *et al* found that vitamin D deficient patients (< 25 ng/mL) are 5.84 times [aOR=6.84, p=0.01] more likely to die from COVID-19 compared to the vitamin D sufficient people (Table 2). Similarly the study by Radujkovic *et al*. found that Vitamin D deficiency is associated with higher risk of death with Hazard ratio (HR) 14.73, p<0.001.

**Table 2:** Findings from the studies.

| Author | Main findings |
| --- | --- |
| Abrishami *et al.*, 2020 | • The probability of death in patients with vitamin D deficiency [defined as 25(OH)D concentration < 25 ng/mL] was 34.6% compared with 6.4% in patients with sufficient vitamin D levels ($P = 0.003$).<br>• Odds of death was significantly higher in vitamin D deficient patients (< 25 ng/mL) [aOR=6.84, p=0.01] in comparison with discharged patients. |
| Maghbooli *et al.*, 2020 | • Severe disease infection was more prevalent in vitamin D deficiency patients compared to vitamin D sufficiency patients (77.2% vs 63.6%) |
|  | patients who had a 25(OH)D<30 ng/mL that is vitamin D deficient had more risk [RR=1.59, p=0.02] of having severe disease infection compared to the patients who had 25(OH)D>=30 ng/mL that is vitamin D sufficient. |
| Radujkovic *et al.*, 2020 | Vitamin D deficiency was associated with higher risk of death (HR= 14.73, p=<0.001) |
| Hastie *et al.*, 2020 | Vitamin D deficiency univariably has significant effect on COVID-19 infection which is deficient people are more likely to be positive [OR=1.37, p=0.011]. But vitamin D deficiency has no significant impact on COVID-19 infection after adjusting for confounders. |
| Otros *et al.*, 2020 | After adjusting for age, gender, obesity, and severe CKD, the odds ratio for Vitamin D deficiency was 3.2 (95 % CI: 0.9-11.4), p = 0.070 |
| Merzon *et al.*, 2020 | The mean plasma vitamin D level was significantly lower among those who tested positive than negative for COVID-19 [19.00 ng/mL vs. 20.55]<br>After controlling for the confounders, patients with low 25(OH)D (<30 ng/mL) level were more likely [aOR=1.45, |
|  | p=<0.001] to be COVID-19 infected compared to the patients with 25(OH)D level $\geq 30$ ng/mL. |
| Meltzer *et al.*, 2020 | Patients with likely deficient vitamin D status at the time of COVID-19 testing had an increased relative risk of testing positive for COVID-19 (relative risk, 1.77; 95%CI, 1.12-2.81; $P = .02$) compared with patients with likely sufficient status at the time of COVID-19 testing, for an estimated mean rate in the deficient group of 21.6% vs 12.2% in the sufficient group |
| Ye *et al.*, 2020 | The serum 25(OH)D levels in COVID-19 patients (55.6 nmol/L) were statistically lower than in healthy controls (71.8 nmol/L)<br>Serum 25(OH)D levels in severe/critical COVID-19 cases (38.2 nmol/L) were significantly lower than that in mild/moderate cases (56.6 nmol/L) |
| Kaufman *et al.*, 2020 | The SARS-CoV-2 positivity rate was lower in the 27,870 patients with “adequate” 25(OH)D values (30–34 ng/mL) (8.1%), than in the 39,190 patients with “deficiency” (<20 ng/mL) (12.5%) (difference 35%; $p < 0.001$). Similarly, the SARS-CoV-2 positivity rate was lower in the 12,321 patients with 25(OH)D values $\geq 55$ ng/mL (5.9%) than in patients with |
| | adequate values (difference 27%; $p < 0.001$) |
| D'avolio *et al.*, 2020 | Observed statistically significant ($p = 0.004$) lower 25(OH)D levels (11.1 ng/mL) in patients positive for the SARS-CoV-2 PCR compared with the negative patients (24.6 ng/mL) |
| Baktash *et al.*, 2020 | Vitamin D levels in the COVID-19-positive group were overall significantly lower compared with that in the COVID-19-negative group (27.00 nmol/L vs 52.00 nmol/L) ($p = 0.0008$) |

The result of the study by Maghbooli *et al* says that among the COVID-19 patients, vitamin D deficient (<30 ng/mL) patients have more risk of developing severe infection compared to the vitamin D sufficient (>*=*30 ng/mL) patients. Similarly, the study by Otros *et al*. found that vitamin D deficient (<12 ng/mL) people are 2.2 times [aOR=3.2, p=0.07] more likely to develop severe COVID-19 after adjusting for age, gender, obesity, cardiac disease, and kidney disease compare to the vitamin D sufficient people. Furthermore, the study by Ye *et al*. found that Serum 25(OH)D levels in severe/critical COVID-19 cases (38.2 nmol/L) are significantly lower than that in mild/moderate cases (56.6 nmol/L).

The study by Merzon *et al*. found that after controlling for confounders, patients with low 25(OH)D (<30 ng/mL) level are more likely [aOR=1.45, p=<0.001] to be COVID-19 infected compare to the patients with 25(OH)D level >=30 ng/mL. Meltzer *et al*. found that patients with likely deficient vitamin D status at the time of COVID-19 testing have more risk of testing positive for COVID-19 (RR= 1.77; 95%CI, 1.12-2.81; *p* = .02) compared to the patients with likely sufficient status at the time of COVID-19 testing. Similarly, Kaufman *et al*. found that the SARS-CoV-2 positivity rate is lower in patients with “adequate” 25(OH) D values (30–34 ng/mL), than in patients with “deficiency” (<20 ng/mL) (difference 35%; p<0.001). The study by D’avolio *et al*. observed significantly lower 25(OH) D levels (11.1 ng/mL) in patients positive for the SARS-CoV-2 PCR compared with the negative patients (24.6 ng/mL). The study by Baktash *et al*. observed that vitamin D levels in the COVID-19-positive group are overall significantly lower compared with that in the COVID-19-negative group. Finally, the study by Hastie *et al*. found that univariably vitamin D deficient people are more likely [OR=1.37, p=0.011] to be COVID-19 infected compare to the vitamin D sufficient people but this relation is not significant in the presence of the confounders. More findings from the included studies are presented in table 2.

## Discussion

The striking relationship between Vitamin D deficiency and COVID-19 risk factors such as obesity and older age has influenced some scholars and researchers to postulate that Vitamin D supplements could be used as a preventive or protective agent against COVID-19 infection, illness and mortality (Martineau, &Forouhi, 2020). Some researchers and clinicians have also argued that since COVID-19 is associated with immune hyperactivation (Zhong et al., 2020), Vitamin D regulates immunopathalogical inflammatory responses and support innate antiviral effector mechanism (Martineau, &Forouhi, 2020). Zhong et al. (2020) and Panarese, &Shahini (2020) independently report that Vitamin D acts as a protective effect against COVID-19 because it boosts immune system response to the SARS-CoV-2 virus. This has also led to a substantial increase in the use of Vitamin D across the globe since the emergence of COVID-19. Several studies (Baktash et al., 2020; Ye et al., 2020; Kaufman et al., 2020) have investigated the likely protective and mitigating effect of these supplements against COVID-19 infection and mortality. We are contributing in this direction by performing this first systematic review of existing evidence on the effect of Vitamin D on COVID-19 status. In all, a total of 11 studies were included in the final analysis. Evidence in all the studies except Hastie and colleagues’ study suggest that Vitamin D has a protective or mitigating effect against COVID-19, either by reducing the risk of contracting, becoming severely ill or dying from COVID-19. In other words, patients who are Vitamin D sufficient are less likely to be infected or critically ill or die from the COVID-19 and vice versa.

The uniformity of evidence in the included studies in our systematic review suggest that there is a possibility that Vitamin D supplementation might reduce the impact of COVID-19 especially in patients and populations with high prevalence of vitamin D deficiency. It may also explain the global increase in demand and utilization of food supplements especially Vitamin D across healthcare and non-healthcare settings amid the COVID-19 pandemic (Ebadi, &Montano-Loza, 2020; Grant et al., 2020; McCartney, &Byrne, 2020). Though the evidence from the review is positive and interesting, our discussion from the systematic review is driven from the relative lack of scientific knowledge relating to biological explanation for the Vitamin D impacts on COVID-19 status since only studies reporting likelihood were available and included. The studies reviewed and included in the evidence synthesis are based on cross-sectional, observational and prospective data and they do not provide indication of the relevant causative agents and mechanisms through which Vitamin D serves as a protective or mitigating effect against COVID-19. This limitation does not suggest that the evidence of positive association between Vitamin D and COVID-19 status must be dismissed.

However, without indicating the causative agents and plausible mechanism in the included studies, the level of confidence of the Vitamin D impact on COVID-19 status is moderate. This is because if a COVID-19 patient is on Vitamin D, a number of factors will determine whether a good effect is likely to occur. These factors will include dose, duration, age, gender, diet, lifestyle state, state of health, among others. These confounding factors may prove important in the Vitamin D and COVID-19 status association. Evidently, one of the studies selected and included in this review (Hastie et al., 2020) show some uncertainties and also suggest that confounders should be taken into consideration when discussing effect of Vitamin D on COVID-19 status. The authors found that Vitamin D does not have significant relationship with COVID-19 status in the presence of confounders. According to Martineau, and Forouhi (2020), results from studies investigating the potential and actual impact of Vitamin D on COVID-19 status appear conflicting to date partly due to the evidence that those studies are open to residual and unmeasured confounding.

From this perspective, epidemiological and hospital-based studies, and well-designed and powered randomised controlled trials are currently required to establish the actual effect of Vitamin D on COVID-19 status and to also offer insights into the potential for reverse causality (Martineau, &Forouhi, 2020). The design of such studies and trials should be informed by findings of systematic reviews on Vitamin D effect on COVID-19 status such as the present review and meta-analyses of randomised controlled trials of vitamin D.

## Conclusion

Findings from the study included suggest Vitamin D may serve as a mitigating effect for covid-19 infection, severity and mortality. We recommend the need to encourage people to eat foods rich in vitamin D such as fish, red meat, liver and egg yolks whiles at the same time providing vitamin D supplements for individuals with COVID-19 in order to boost their immune systems.

## Data Availability

Data is available upon request

## Authors’ contributions

PD developed the study concept, developed search strategy, conducted literature search, quality assessed and extracted data. PD, NS, BOA, PP, AM, AS drafted the manuscript. And critical review by AS.

## Acknowledgements

Not applicable.

## Consent for publication

Not applicable.

## Ethics approval and consent to participate

Not applicable

## Availability of data and materials

The data used to support the findings of this study are available from the corresponding author upon request.

