## Supplementary file for "Effect of Vitamin D deficiency on COVID-19 status: A systematic review"

**Supplementary tables**

**Table S1**: PubMed search results for Vitamin D status and COVID-19.

| **Search strategy** | **Results** |
| --- | --- |
| ("Vitamin D," OR "25‐Hydroxyvitamin D," OR "Low Vitamin D.") AND ("COVID-19" OR "2019-nCoV" OR "Coronavirus" OR "SARS-CoV-2") AND ("disease severity" OR "IMV" OR "ICU admission" OR "mortality" OR "hospitalization" OR "infection")  **Timespan**: 1/12/2019 to 3/11/2020, **Article type**: Journal article, **Language**: English | 135 |

**Table S2:** Google Scholar search results for Vitamin D status and COVID-19.

| **Search strategy** | **Results** |
| --- | --- |
| ("Vitamin D," OR "25‐Hydroxyvitamin D," OR "Low Vitamin D.") AND ("COVID-19" OR "2019-nCoV" OR "Coronavirus" OR "SARS-CoV-2") AND ("disease severity" OR "IMV" OR "ICU admission" OR "mortality" OR "hospitalization" OR "infection")  **Timespan**: Since 2020 **Language**: English | 2630 |

**Table S3:** ScienceDirect search results for Vitamin D status and COVID-19.

| **Search Strategy** | **Results** |
| --- | --- |
| ("Vitamin D," OR "25‐Hydroxyvitamin D," OR "Low Vitamin D.") AND ("COVID-19" OR "2019-nCoV" OR "Coronavirus" OR "SARS-CoV-2") AND ("disease severity" OR "IMV" OR "ICU admission" OR "mortality" OR "hospitalization" OR "infection")  **Article type:** Research article | 9 |

**Table S4.** Newcastle-Ottawa scale assessment of study quality for **cross-sectional study**

| Author | Selection | | | | Comparability | Outcome | | Study  quality |
| --- | --- | --- | --- | --- | --- | --- | --- | --- |
|  | 1 | 2 | 3 | 4 | 5 | 6 | 7 |  |
|  | Representativeness of the sample | Sample size | Ascertainment of exposure | Non-respondents | The subjects in different outcome groups are comparable, based on the study design or analysis. Confounding factors are controlled. | Assessment of outcome | Statistical test is appropriate |  |
| Maghbooli *et al.*, 2020 | * | * | * |  | * | * | * | 6 |
| Hastie *et al.*, 2020 | * | * | * |  |  | * | * | 5 |
| Merzon *et al.*, 2020 | * | * | * |  |  | * | * | 5 |
| Kaufman *et al.*, 2020 | * | * | * | * |  | * | * | 6 |

**Table S5.** Newcastle-Ottawa scale assessment of study quality for **cohort study**

| Author | Selection | | | | Comparability | | Outcome | | | Study Quality |
| --- | --- | --- | --- | --- | --- | --- | --- | --- | --- | --- |
|  | 1 | 2 | 3 | 4 | 5A | 5B | 6 | 7 | 8 |  |
|  | Exposed cohort truly representative | Non-exposed cohort drawn from the same community | Ascertainment of exposure | Outcome of interest not present at start | Cohorts comparable on basis of age | Cohorts comparable on other factor(s) | Quality of outcome assessment | Follow-up long enough for outcomes to occur | Complete accounting for cohorts |  |
| Abrishami *et al.*, 2020 | * |  | * | * |  |  | * | * |  | 5 |
| Radujkovic *et al.*, 2020 | * |  | * | * |  |  | * | * |  | 5 |
| Otros *et al.*, 2020 | * |  | * | * |  |  | * | * |  | 5 |
| Meltzer *et al.*, 2020 | * |  | * | * |  |  | * | * |  | 5 |
| D’avolio *et al.*, 2020 | * | * | * | * |  |  | * | * |  | 6 |
| Baktash *et al.*, 2020 | * |  | * | * |  |  | * | * |  | 5 |

**Table S6.** Newcastle-Ottawa scale assessment of study quality for **case control study**

| Author | Selection | | | | Comparability | Outcome | | | Study quality |
| --- | --- | --- | --- | --- | --- | --- | --- | --- | --- |
|  | 1 | 2 | 3 | 4 | 5 | 6 | 7 | 8 |  |
|  | Is the case definition adequate? | Representativeness of the cases | Selection of Controls | Definition of Controls | Comparability of cases and controls on the basis of the design or analysis | Ascertainment of exposure | Same method of ascertainment for cases and controls | Non-Response rate |  |
| Ye *et al.*, 2020 | * | * | * | * | * | * | * |  | 7 |
